# Very Long-term Longitudinal Follow-up of Heart Failure on the REMADHE Trial

**DOI:** 10.1101/2024.03.26.24304939

**Authors:** Edimar Alcides Bocchi, Guilherme Veiga Guimaraes, Cristhian Espinoza Romero, Silvia Moreira Ayub Ferreira, Bruno Biselli, Paulo Roberto Chizzola, Robinson Tadeu Munhoz, Julia Tizue Fukushima, Fátima das Dores Cruz

**Author notes:** **Correspondence Address:** Prof Dr Edimar Alcides Bocchi, Rua Dr Melo Alves no 690, apto 41. São Paulo, Brazil, CEP 01417-010.

## Abstract

**Background:** Heart failure (HF) is associated with frequent hospitalization and worse prognosis. Prognosis factors and survival in very long-term follow-up have not been reported in HF. HF disease management programs(DMP) results are contradictory. DMP efficacy in very long-term follow-up is unknown. We studied the very long-term follow-up of up to 23.6 years and prognostic factors of HF in 412 patients under GDMT included in the REMADHE trial.

**Methods:** The REMADHE trial was a prospective, single-center, randomized trial comparing DMP versus usual care(C). The first patient was randomized on October 5, 1999. The primary outcome of this extended REMADHE was all-cause mortality.

**Results:** The all-cause mortality rate was 88.3%. HF was the first cause of death followed by death at home. Mortality was higher in the first 6-year follow-up. The predictive variables in multivariate analysis associated with mortality were age ≥52 years (P=0.015), Chagas etiology (P=0.010), LVEF <45% (P=0.008), use of digoxin (P=0.002), functional class IV (P=0.01), increase in urea (P=0.03), and reduction of lymphocytes (P=0.005). In very long-term follow-up, DMP did not affect mortality in patients under GDMT. HF as a cause of death was more frequent in the C group. Chagas disease, LVEF <45%, and renal function were associated with different modes of death.

**Conclusion:** DMP was not effective in reducing very-long term mortality; however, the causes of death had changed. Our findings that age, LVEF, Chagas’ disease, functional class, renal function, lymphocytes, and digoxin use were associated with poor prognosis could influence future strategies to improve HF management.

---

Heart failure (HF) has an estimated prevalence of 1 to 4% of the global population.^1^ HF remains associated with poor quality of life, high mortality, hospitalizations, and is a substantial burden on the healthcare system. HF may have heterogeneous causes and pathways. However, HF trials and observational survival studies conducted worldwide have relatively limited short follow-up periods.^2–4^ Few studies have assessed the long-term survival impact of HF beyond a 10-year period.^5–11^ Thus, data regarding very long-term survival and respective prognostic factors in HF are lacking.

Educational and disease management programs (DMP) targeted at patients with HF have reported improvement in quality of life, and reduction in hospitalization and healthcare utilization.^11–13^ However, doubt has been cast on the efficacy of these interventions based on several published neutral studies.^14^, In fact, very long-term efficacy of DMP in HF is unknown.

The REMADHE trial was conducted initially during a mean follow-up period of 2.47±1.75 years. The study demonstrated improvements in quality of life, reductions in hospitalizations and emergency visits among the DMP group, without statistical differences in mortality rates between the groups.^13^ The objective of our current study was to extend the follow-up period of patients initially included in the REMADHE trial up to 23.6 years. Also, we aimed to identify prognostic predictors of all-cause mortality in a population with HF who initially underwent education and telephone monitoring in a specialized and multidisciplinary HF unit.

## Methods

REMADHE was a prospective, single center, open trial with randomization 2:1, as previously detailed.^13^ The REMADHE study compared the DMP group versus the Control group (C), in patients treated in a clinic specializing in HF with a multidisciplinary team. Patients in the DMP group underwent an educational program and continuous repetitive monitoring. Patients received reinforcement of education during the 2.47±1.75-year follow-up at 6-month intervals. Education and monitoring were not repeated with frequent reinforcement throughout the very late follow-up. In this current extended study, we analyzed on June 2023 data of patients included in the period from October 1999 to January 18, 2005, with follow-up until 23.6 years.

Data about death were obtained from reports collected during medical visits, telephone calls, review of medical records, information from family members on data contained in the death certificate, research at the SEADE Foundation (State Data Analysis System), and the central deaths in Brazil (Ministry of Health).

### Study population

The patients included in the very long-term follow-up of the REMADHE trial were initially recruited from a tertiary cardiological referral center who were undergoing outpatient follow-up with cardiologists specialized in heart failure (HF) at the Heart Failure Clinics. All patients were under guideline-directed medical therapy (GDMT). The eligibility criteria and exclusion criteria have been described previously.^13^

### Statistical analysis

Descriptive statistics of quantitative variables were performed using mean (M), standard deviation (SD), and number of cases (N). Relative variations (Δ%) were also calculated and, if this was not possible, absolute variations (Δ) were evaluated between the results of the sequential follow-up of each variable. The distribution of quantitative variables was evaluated using the Kolmogorov-Smirnov test. Categorical variables were described with absolute and relative frequencies. Normality was determined by the Shapiro-Wilk test. The Student *t* test was used to compare the baseline characteristics of groups C and DMP, and the Fisher exact test was used for unpaired values. In the analysis of mortality, the date of randomization up to the data obtained by telephone, by medical records or by death certificate was considered. Survival and event-free curves were calculated using the Kaplan-Meier method, and the log-rank test (Mantel-Cox) was used for comparison. P<0.05 was considered statistically significant.

A univariate and multivariate proportional hazards model was adjusted to assess prognostic factors associated with mortality outcome. The following variables were tested initially on a univariate model: sex, age < or ≥52 years, ethnicity (white, black, mulatto), etiology (ischemic, hypertensive, alcoholic, chagasic, valvular, and others), diabetes type II, diabetes insulin-dependent, left ventricular ejection fraction (LVEF) ≥ or <45%, left bundle-branch block, implanted pacemaker, digoxin use, New York Heart Association function class (functional class), education level, marital status, quality of life (Minnesota Questionnaire), blood plasma levels of sodium, potassium, urea, creatinine, glycemia, hemoglobin, leucocytes, thyroid hormones (T3/T4), thyroid stimulating hormone, and uric acid. Variables with P<0.10 values were used to compose the multivariate model with a stepwise variable selection process. P values <0.05 were considered significant. A baseline characteristic analysis was conducted to investigate potential confounding factors among the positive predictor variables examined in the multivariable analysis. Statistical analysis was performed with SPSS v 21 (SPSS Inc, Chicago, IL).

## RESULTS

### Demographic Data

Groups DMP and C had similar demographic baseline characteristics, with a total of 412 included patients as previously published.^15^ The time between the first randomization and outcome analysis was 23.6 years. The baseline characteristics of the patients were previously published in the initial study.^13^

### Total Mortality

Mortality data were analyzed from October 1999 to June 2023, showing all-cause mortality rate of 88.3% (**Fig. 1A**). HF was the cause of death in 35.9% (n=132) of patients who died; 25.5% (n=105) died at home, other causes of death were observed in 19.3% (n=79), and in 11.2% (n=46) the cause was unknown. The inclination of the survival curve is higher in the first 6-year follow-up in comparison with after 6-year follow-up (**Fig. 1A**). The survival curves according to DMP and C groups are shown in **Fig. 1B**. At 23.6-year follow-up, univariate analysis revealed that several variables were associated with lower survival rates (**Table 1**), including age ≥52 years (**Fig. 2A**), LVEF <45% (**Fig. 2B**), chagasic etiology (**Fig. 2C**), digoxin users (**Fig. 2D**), urea levels (**Fig. 3A**), lymphocytes (**Fig. 3B**), and functional class IV (**Fig. 3C**), male sex **(Fig. 4A)**, and atrial fibrillation (AF) **(Fig. 4B)**. On the multivariate analysis, the predictive variables for mortality were age ≥52 years (HR 1.315; 95% Confidence Interval [CI], 1.055 to 1.640; P=0.015); Chagas etiology (HR 1.672; 95% CI, 1.252 to 2.232; P<0.001); LVEF <45% (HR 0.582; 95% CI, 0.389 to 0.870; P=0.008); use of digoxin (HR 1.425; 95% CI, 1.138 to 1.785; P=0.002); functional class IV (HR 1.604; 95% CI, 1.122 to -2.295; P=0.010); elevation of urea levels (HR 1.008; 95% CI, 1.003 to 1.014; P=0.038); and reduction of lymphocytes (HR 0.772; 95% CI, 0.641 to 0.929; P=0.005).

**Figure 1.**
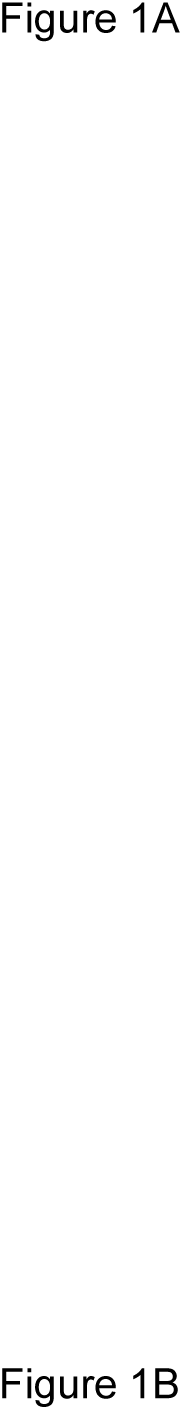
Kaplan-Meier Survival Curve in the Total Population (Figure 1A). The Survival Curves According to Intervention and Usual Care Groups (Figure 1B).

**Table 1.**
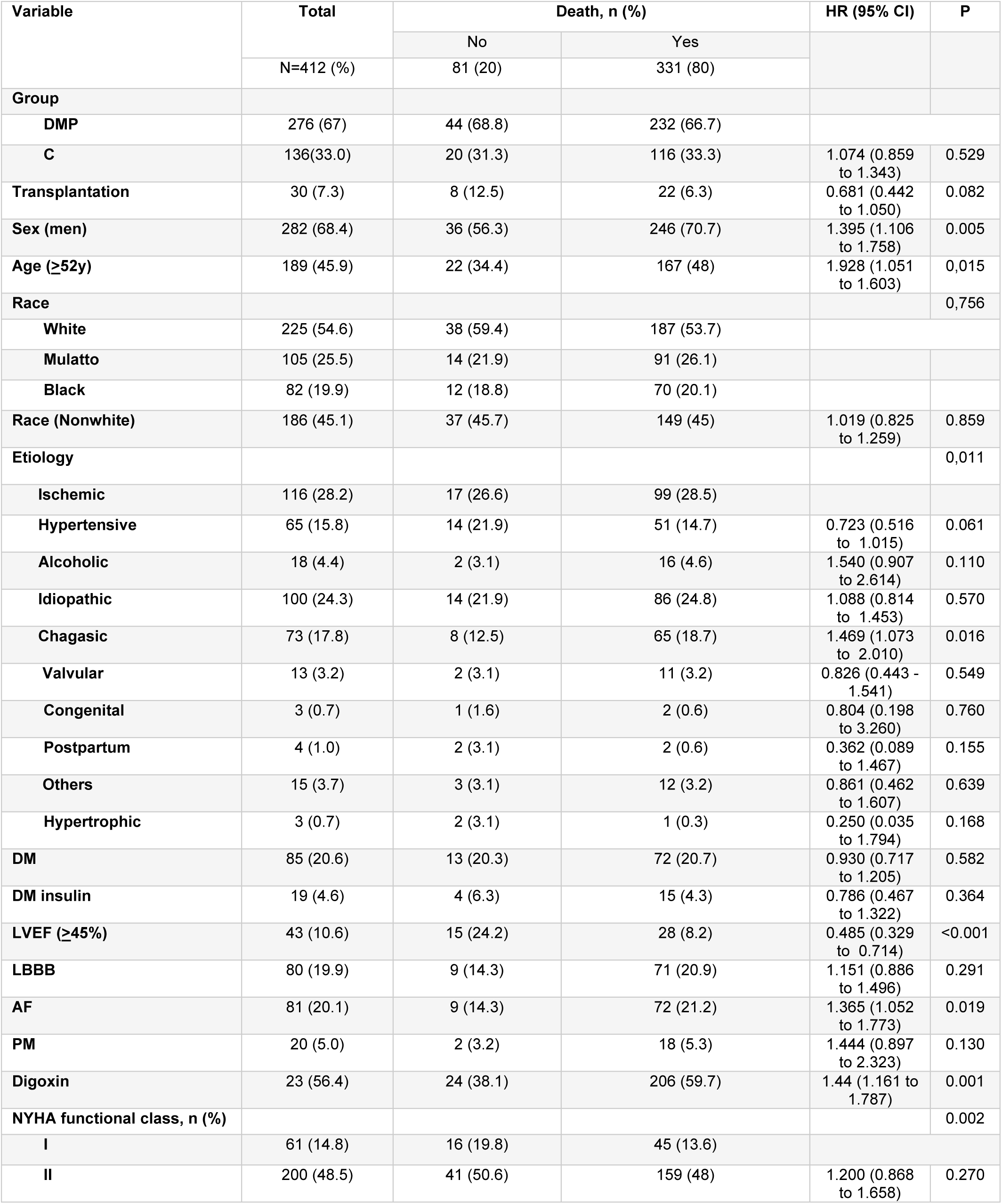
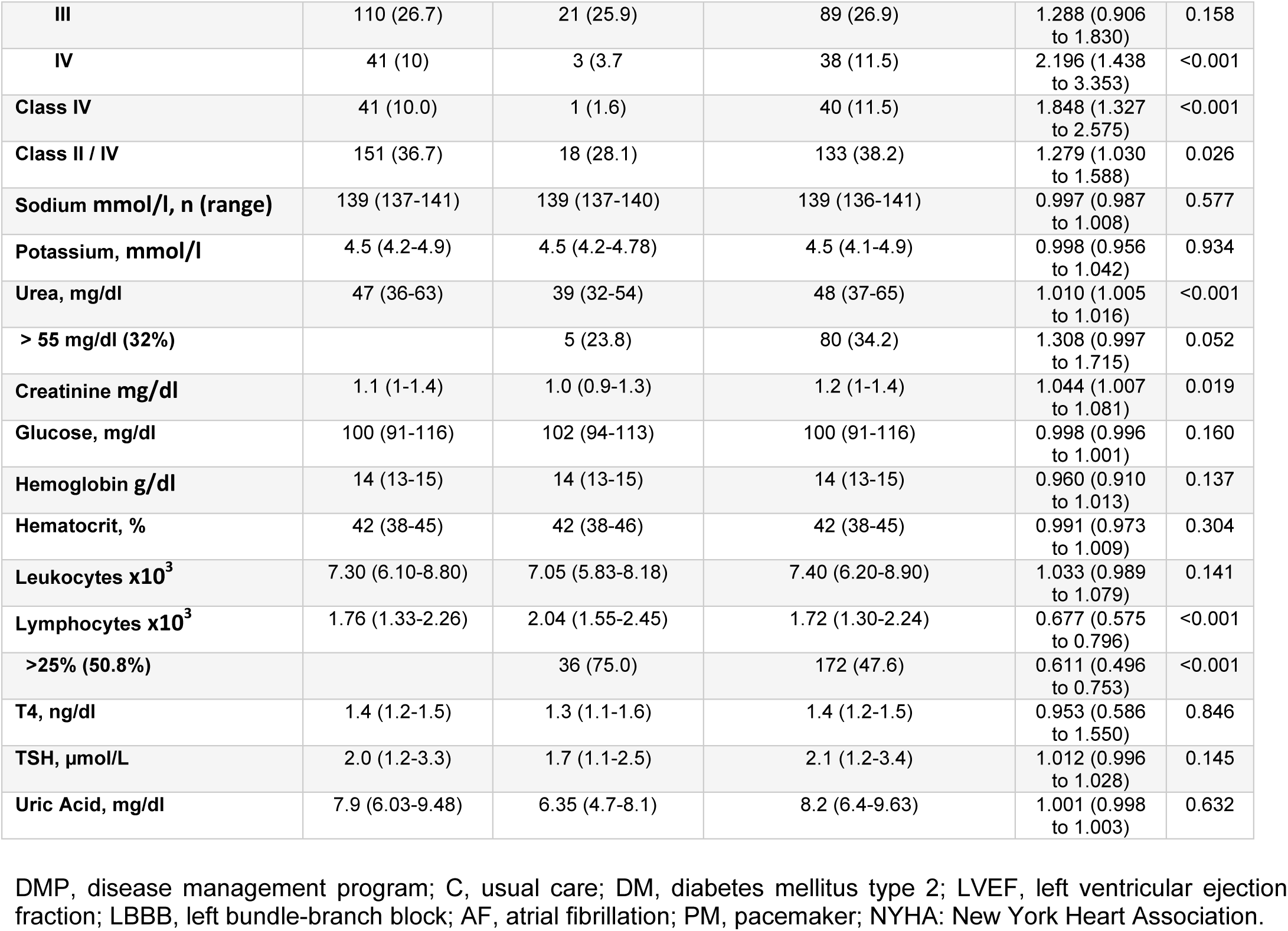
Univariable Analysis of Predictors Associated With Any Mortality at 23.7-year follow-up.

**Figure 2.**
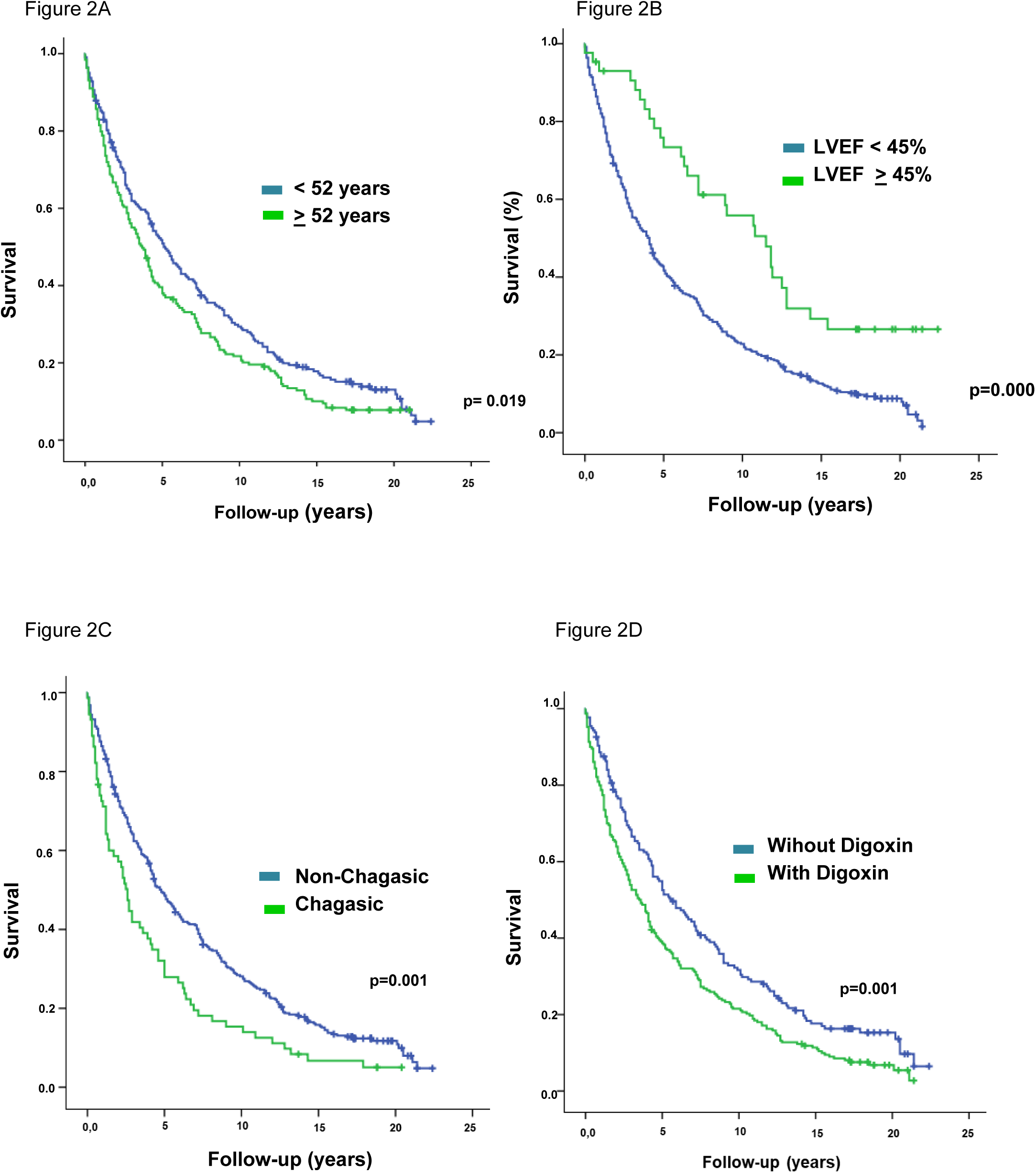
Kaplan-Meier Survival Curve in Subgroup Analysis; 2A, According to Age <52 and *≥*52 Years; 2B, According to Left Ventricular Ejection Fraction <45% and ≥45%; 2C, According to Chagas’ and non-Chagas’ Etiology; 2D, According to Digoxin Use.

**Figure 3.**
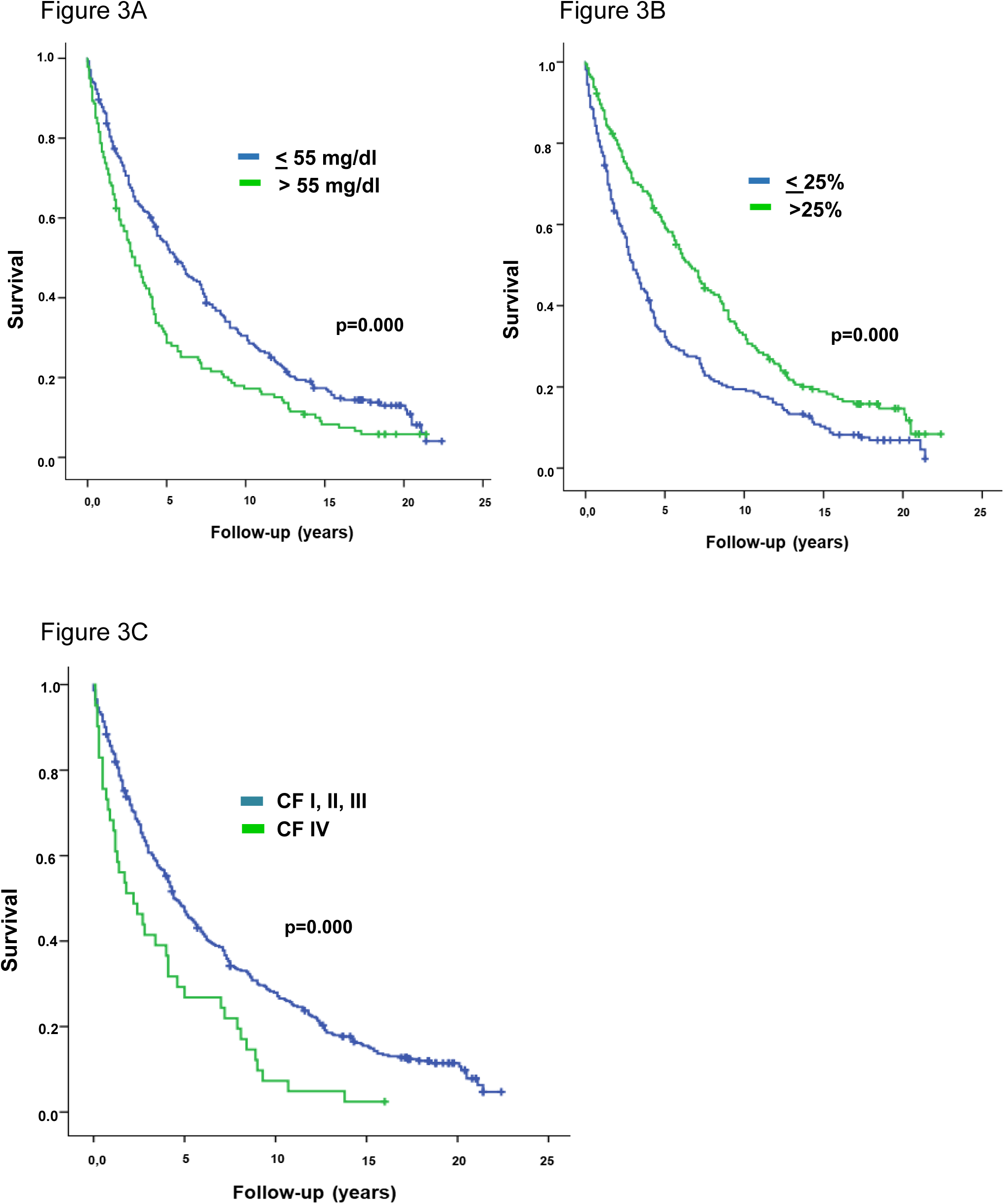
Kaplan-Meier Survival According to Subgroup Analysis: 3A, According to Urea Levels ≤55 mg/dl and >55 mg/dl; 3B, According percentage of Lymphocytes >25% versus ≤25%; 3C, According New York Association Functional Class

**Figure 4.**
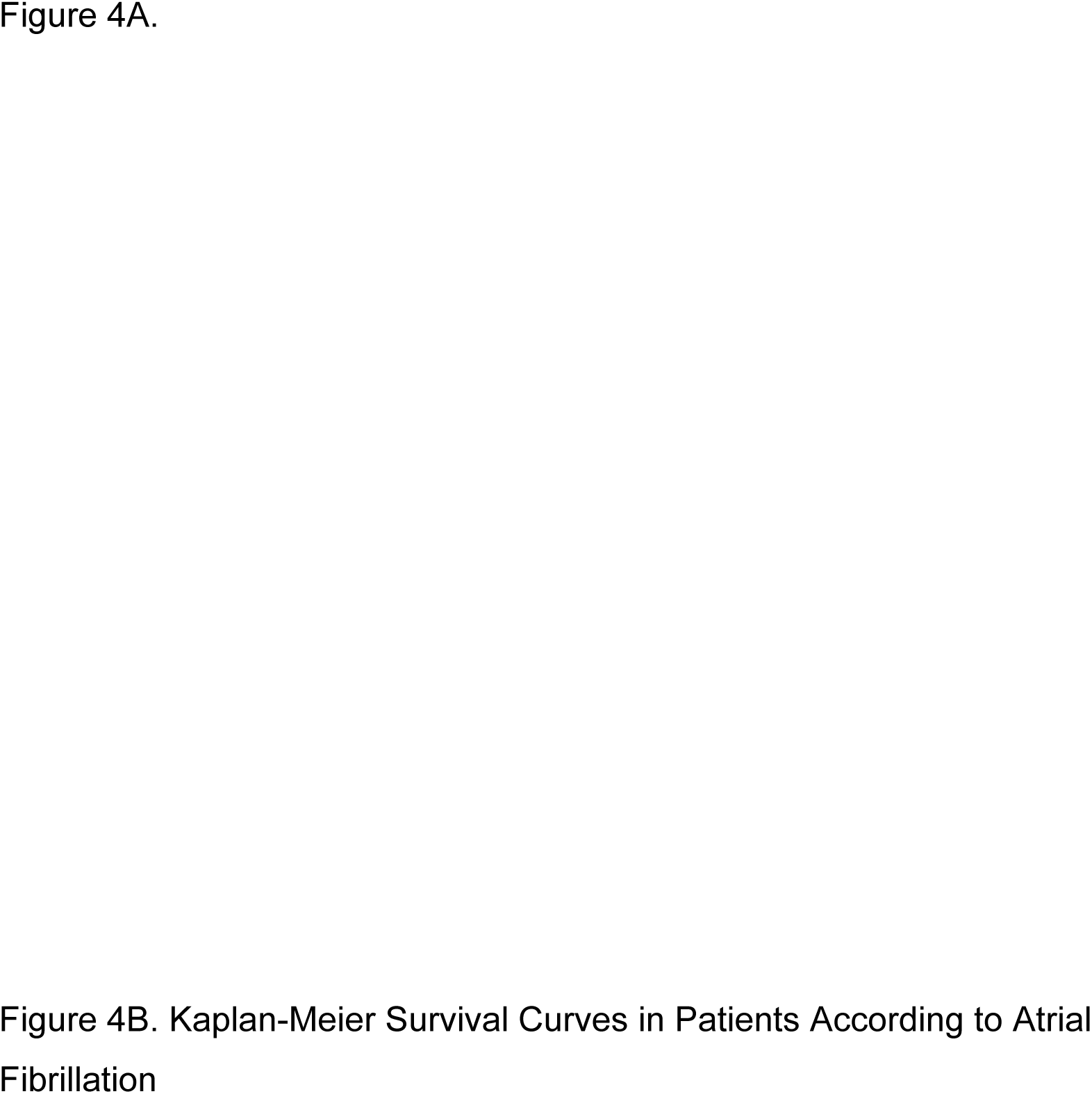
Kaplan-Meier Survival Curve:4A According Sex Men and Woman, 4B and Kaplan-Meier Survival Curve According Patients With Atrial Fibrillation

Chagas disease tended to differ in causes of death compared with other etiologies (P>0.07). In death from Chagas’ disease, HF was the cause in 43.2%, death at home was observed in 17.6%, other causes in 33.8%, and unknown in 5.4%. In non-Chagas’ disease deaths, HF was the cause in 37%, death at home was observed in 34.6, other causes in 17.1, and unknown in 11.3%. Causes of death were different according to baseline LVEF <45% and ≥45% (P<0.04). In LVEF < 45% HF as a cause of death, death at home, other causes, and unknown causes were 40, 29.7%, 21.7%, and 8.6 %, respectively. In LVEF ≥45%, HF cause of death, death at home, other causes, and unknown causes were 16.7%, 29.7%, 21.7%, and unknown in 8.6 % respectively. Causes of death were different according to baseline urea >55 mg/dl and urea ≤55 mg/dl (P<0.01). In urea >55mg/dl, HF as the cause of death, death at home, other causes, and unknown causes were 46%, 24.2%, 18.5%, and 11.3%, respectively. In urea ≤55%, HF as the cause of death, death at home, other causes, and unknown causes were 34%, 35%, 21.8%, and 9.2%, respectively. Other independent variables related to mortality in multivariate analysis did not influence the causes of death.

### Mortality between groups

The mean survival was 6.2±0.52 years in C versus 6.6±0.37 years in DMP (P=0.656) up to 23.6-year follow-up (**Fig. 1B**). HF as a cause of death and death at home were different between groups DMP and C (P<0.02). HF during hospitalization was the cause of death in 33.3% and 41% in DMP and C groups, respectively; and death at home was observed in 28.4% and 20.4% of deaths in DMP and C groups, respectively. Other causes of death or unknown causes were observed in 34.7%, and 34.2% of the deaths in DMP and C groups, respectively (P=ns).

## Discussion

One of the notable strengths of this study is the very long-term follow-up of patients with HF, which, to the best of our knowledge, represents the first DMP analysis of a follow-up period exceeding 20 years. The main findings can be summarized as follows: (1) the survival of HF patients analyzed over a 20-year period showed during the first 6 years an inclination of the survival curve suggesting initial high risk even in patients under ambulatory care; (2) age (≥52 years), Chagas disease, LVEF <45%, use of digoxin, functional class IV, elevated urea levels, and lower percentage of lymphocytes were independent predictors of mortality; (3) DMP and C groups had similar survival. However, HF as cause of death was more frequent in C; (4) HF was the first cause of death followed by death at home; (5) Some independent variables on multivariate analysis were associated with different modes of death, including Chagas’ disease; baseline LVEF and renal function.

This study is novel in the analysis of very long-term mortality (exceeding 20 years) in HF patients and who underwent DMP. Our results showed better HF survival in comparison with recently reported HF data up to 10-year follow-up.^5^ One reason for this would be that our patients were followed up by HF specialists in a Heart Failure Clinic. Also, our findings add new data on modes of death in very long-term follow-up on HF. Mechanisms related to higher mortality for approximately the first 6 years are unknown. The higher mortality up to 5 years was also reported recently after HF hospitalization. Those who responded poorly or not at all to triple therapy including those who did not maintain the initial response could have died in the first years instead of those who responded to therapy and had longer follow-up. Patient characteristics under optimized therapy as observed in our results could influence the response to treatment. As main implications of our results independent modifiable markers with a strong pathophysiological rationale could be priority targets for treating or planning research on HF in very long-term follow-up. Renal function and LVEF seem to have these characteristics.

One hypothesis to explain the no effects of REMADHE DMP on very long-term mortality might be the continuous GDMT in both DMP and C groups in patients followed in a Heart Failure Clinic. Patients included in the REMADHE study received ongoing specialized care at the HF unit throughout the entire extended duration of the study. However, the finding that the C group was associated with HF as a more important cause of death suggests the DMP may be effective in preventing events related to progressive HF, but not in preventing in-home death presumably most due to sudden death. Mechanisms related to sudden death in HF are multiple and complex.^15^ Prevention of events associated with progressive HF may have driven sudden death in very long-term follow-up.

Worse prognosis of chagasic HF on shorter follow-up was also observed in the extended long-term follow-up.^13^ Despite already being first described in 1994 by Bocchi et al, the mechanisms related to worse prognosis in chagasic HF still is an unresolved issue.^16^ The complex pathogenesis and physiopathology comprising persistent myocarditis with fibrosis, parasite persistence with inflammatory response, autoimmunity, damage to the parasympathetic system causing sympathetic over activity, microvascular abnormalities, conduction system abnormalities, brady- and tachyarrhythmias, biventricular dilated cardiomyopathy, apical aneurysm, thromboembolism, or remodeled ventricles may be related to worse prognosis.^17–18^ Also, only 35.8% of patients with Chagas disease were receiving baseline beta-blocker therapy. However, the lack of knowledge about whether GDMT is effective for chagasic HF may have influenced the smaller proportion of beta-blocker therapy compared with other etiologies. Medical treatment has been extrapolated from trials that included other etiologies or studies with limited design.^17^ However, a subanalysis of the REMADHE trial showed that the survival of patients with Chagas disease undergoing beta-blocker therapy was similar to that of other etiologies.^19^

Our results on multivariate analysis concerning age align with findings of studies that reported a negative impact of aging on survival. ^5^ However, in our cohort, patients were relatively younger (mean age of 51 years), and an age already ≥ 52 years was associated with lower survival. The presence of a younger population can be attributed to earlier manifestation of etiologies such as Chagas’ disease, valvar abnormalities, and limited access to prevention in a population despite risk factors of developed and undeveloped countries.^20^

Our findings on very long-term follow-up are in agreement with prior studies showing that reduced LVEF is a well-established predictor of HF mortality particularly with an average follow-up of up to 5 years.^21–23^ Studies have not explored very longer follow-up periods. Otherwise, heart failure with LVEF > 45% (HFpEF) was also associated with increased mortality mainly in N.Y.H.A functional class IV.^24,25^ However, prognosis of HFpEF is controversial depending of characteristics of included patient in studies. Overall, it is expected patients with recovered LVEF in HFpEF group. Better prognosis was reported in HF with improved LVEF in comparison with persistent HFpEF, declined EF and persistent heart failure with reduced ejection fraction.^21^ Additionally, the worsening of functional class is known to be associated with worse outcomes in HF, which was consistent with our findings in very long-term follow-up. functional class IV was also associated with reduced survival, similar to observations from other studies with follow-up periods of up to 10 years.^24,25^

Concerning the digoxin association with worse prognosis reported in our results, it is crucial to highlight that studies had reported contradictory associations of digoxin with mortality in HF.^26–28^ However, most studies have the caveat of absence of serum digoxin levels assessment, which might have affected outcomes. Subanalysis of the Digitalis Investigation Group trial showed a linear dose–response relationship linking serum concentration to mortality.^29^ Also, the reason for digoxin prescription may be a confounder because in clinical practice digoxin could had been prescribed for more seriously ill patients. The findings emphasize cautious prescribing of digoxin for patients with HF in very long follow-up, because its association with increased mortality was suggested in previous research and by our results. Also, the evidence of benefits of digoxin may be limited in patients undergoing contemporaneous HF treatment.

Our data in which the baseline mean urea values >55 mg/dl were associated with reduced survival confirms previous publications, however, adding new very long-term data. Numerous studies, particularly in the context of decompensated HF, have examined the prognostic value of elevated urea levels (>55-80 mg/dl) as a predictor of morbidity and mortality, albeit with short-term follow-up.^30–31^ Report of the Swedish Heart Failure Kidney Registry showed that renal dysfunction is common and strongly associated with short-term and long-term outcomes up to 10-year follow-up in patients with HF.^31^ Systematic review and meta-analysis reported that worsening renal function predicts substantially higher rates of mortality and hospitalization in patients with HF.^32^

Baseline reduction in the percentage of lymphocytes as a biomarker for prognosis in HF has been reported, but it has not yet been demonstrated in long-term follow-up as in our results.^33–35^ Lymphopenia is also a marker for worse prognosis in other systemic diseases including COVID-19.^36^ The mechanisms responsible for the increment in the relative reduction in lymphocytes in HF are not fully understood. An increase in neutrophil because of systemic inflammation, and lymphopenia caused by elevated cytokines, splanchnic congestion, apoptosis, increased endogenous cortisol and sympathetic tone may play a role.^37,38^ HF can trigger a significant increase in systemic cortisol production.^55^

## Limitations

Our study, being a post hoc analysis, has several limitations owing to its design. However, this extended study reflects a single-center study that allowed GDMT optimization to be maintained.

In conclusion, in a very long-term follow-up exceeding 20 years independent mortality predictors were age ≥52 years, Chagas disease, LVEF<45%, use of digoxin, functional class IV, elevated urea levels, and lower percentage of lymphocytes No significant differences in mortality were observed between the DMP intervention and control groups. However, DMP changed the cause of death, and HF as cause of death was more frequent in the control group. Data our study would be highly valuable for patients, doctors, and healthcare professionals seeking a comprehensive understanding and strategies for management of HF in very long-term follow-up.

## Data Availability

We declare that all data referred to in the manuscript are available upon request to the author.

